# Multidomain Lifestyle Profiles, Biomarker-Defined Alzheimer’s Pathology, and Clinical Expression: Design of the HUNT-ADAPT Study

**DOI:** 10.64898/2026.06.16.26355748

**Authors:** Kevin O’Hara-Veintimilla, Elisabeth Lind Melbye, Miguel German Borda, Poppy AC Mallinson, Anita Lenora Sunde, Antoine Leuzy, Mark van der Giezen, Pier-Giorgio Masci, Yohannes Seyoum, Beatriz Pozuelo Moyano, Felipe Botero-Rodriguez, Michael C. Craig, Lizhi Guo, Lingfeng Xue, Håvard K. Skjellegrind, Ragnhild Oesterhus, Audun Osland Vik-Mo, Diego A. Tovar-Rios, Mira G.P. Zuidgeest, Holger Fröhlich, Chiara de Lucia, Richard Siow, Miia Kivipelto, Ole A. Andreassen, Geir Selbæk, Dag Aarsland

## Abstract

**Background:** It remains unclear why biomarker-defined Alzheimer’s disease neuropathological change (ADNC) leads to cognitive decline and dementia in some individuals but not others. Multidomain lifestyle profiles may influence both pathology risk and clinical expression.

**Objectives:** To determine whether multidomain lifestyle profiles are associated with (1) the risk of biomarker-defined ADNC and (2) the clinical expression of AD pathology, including longitudinal cognitive test score change and incident dementia.

**Design/Setting:** Retrospective longitudinal population-based cohort study within the Trøndelag Health Study (HUNT), Norway, including five waves over a 40-year follow-up period.

**Participants:** The late-life source population comprises 9,956 HUNT4 70+ participants, of whom 5,729 participated in the Ageing in Trøndelag (AiT) follow-up.

**Outcome measurements:** ADNC is operationalized primarily using plasma phosphorylated tau at threonine 217 (p-tau217), with plasma neurofilament light (NfL) available as a complementary marker of neurodegeneration. In HUNT4 70+, participants aged 70 and older had a standardized cognitive diagnostic assessment, which was repeated at AiT four years later.

**Lifestyle Measurements:** Multidomain lifestyle will be defined across five domains: nutrition, physical activity and skeletal muscle health, mental and social health, cardiovascular/metabolic status, and cognitive stimulation. Genetic susceptibility, including *APOE* and genome-wide/polygenic risk measures, and available multi-omics data will be examined as modifiers.

**Results:** Analyses will examine the associations of multidomain lifestyle profiles and domain-specific exposures with both ADNC risk and clinical expression.

**Conclusions:** This study will test whether multidomain lifestyle profiles are associated with both the risk of biomarker-defined ADNC and the clinical expression, informing risk stratification and modifiable prevention in AD.

## 1. Introduction

Dementia is a growing global public health challenge, and Alzheimer’s disease (AD) remains its most common underlying cause [1]. An unresolved question is why similar levels of AD-related pathology are associated with cognitive impairment or dementia in some individuals but relative clinical preservation in others, i.e. resilience and vulnerability. Although AD neuropathological change (ADNC) becomes more common with ageing, it is neither inevitable nor sufficient on its own to determine clinical dementia. Similar levels of pathology may be associated with markedly different clinical trajectories [2], suggesting that the transition from ADNC to cognitive impairment is shaped by factors beyond pathology alone [3]. Modifiable lifestyle-related factors, genetic susceptibility, comorbidities, sex and reproductive ageing, and reserve-related mechanisms may influence both vulnerability to ADNC and whether, or how, ADNC becomes clinically expressed.

Blood-based biomarkers allow ADNC to be studied at scale [4–7]. Understanding resilience and vulnerability is critical for dementia prevention and for devising innovative treatments. Most prevention frameworks emphasize modifiable risk and protective factors across the life course, but have focused mainly on dementia as a clinical endpoint rather than on separating associations with biomarker-defined AD pathology from associations with its clinical expression. Recent work has shown that all 2024 Lancet Commission modifiable dementia risk factors can be operationalized within a single population-based cohort, with more than half of dementia cases estimated to be attributable to these factors [8, 9]. However, this does not clarify whether multidomain lifestyle profiles relate to ADNC risk, clinical vulnerability among individuals with prevalent ADNC, or both [10].

The Finnish Geriatric Intervention Study to Prevent Cognitive Impairment and Disability (FINGER) provide a practical multidomain prevention framework, showing that coordinated lifestyle intervention can slow cognitive decline in at-risk older adults [11]. Within observational cohorts, examining FINGER-aligned domains may also provide a useful structure for evaluating multidomain lifestyle profiles across the life course. However, multidomain prevention research and biomarker-based AD epidemiology are rarely integrated within a life-course population study [12]. To our knowledge, no large-scale population-based study with repeated life-course exposure assessments has examined whether FINGER-aligned lifestyle profiles relate to both later-life ADNC risk and the clinical expression of ADNC once present [13].

The Trøndelag Health Study (HUNT) offers a unique setting to address this gap [14], with its large population-based cohort, repeated adult-life assessments from age 20 years onward, extensive clinical and questionnaire data, biobank resources, multi-omics data, and registry linkage potential. In HUNT4 70+ and the four-year Ageing in Trøndelag (AiT) follow-up, plasma phosphorylated tau at threonine 217 (p-tau217) and plasma neurofilament light chain (NfL) are available alongside standardized cognitive and clinical assessments in adults aged 70 years and older [15]. The HUNT-ADAPT study—Alzheimer’s Disease, Lifestyle, and Pathology-related Trajectories—is designed to examine whether multidomain lifestyle profiles are associated with biomarker-defined ADNC and whether these profiles modify its clinical expression, i.e. longitudinal cognitive test score change and incident dementia.

### 1.1. Study objectives

The objective of the HUNT-ADAPT study is to determine whether multidomain lifestyle profiles, operationalized through FINGER-aligned domain scores, are associated with the risk of biomarker-defined ADNC and with the clinical expression of Alzheimer’s pathology, including longitudinal cognitive test score change and incident dementia, in the HUNT population.

Specific objectives are:

(1) to operationalize FINGER-aligned multidomain lifestyle domains within HUNT, including nutrition, physical activity and skeletal muscle health, mental and social health, cardiovascular/metabolic status, and cognitive stimulation;
(2) to examine whether the multidomain lifestyle profile and individual FINGER-aligned domains are associated with the risk of biomarker-defined ADNC, operationalized using plasma p-tau217;
(3) to examine whether the multidomain lifestyle profile and individual FINGER-aligned domains are associated with longitudinal cognitive test score change and incident dementia;
(4) to determine whether multidomain lifestyle profiles and individual FINGER-aligned domains modify the association between biomarker-defined ADNC and cognitive test score change or incident dementia;
(5) to examine *APOE* and genome-wide risk measures as modifiers of associations between FINGER-aligned lifestyle profiles, ADNC risk, and pathology-related clinical outcomes;
(6) to evaluate available multi-omics data in relation to ADNC risk, lifestyle-related biological signatures, and their potential modifying role in pathology-related clinical outcomes; and
(7) to examine associations of oral health, reproductive and endocrine ageing indicators with ADNC risk and pathology-related clinical outcomes, including their potential modifying role.

## 2. Methods

### 2.1. Study design and setting

HUNT-ADAPT is a population-based cohort study nested within The HUNT study, a large longitudinal health study conducted in Trøndelag County, Norway. Since 1984, HUNT has invited all adult residents of the region to repeat population-based health surveys approximately every 10 years, integrating questionnaire data, clinical examinations, biological and genetic samples, and registry-linkage capacity through the Norwegian personal identification number system. Across four survey waves and multiple sub studies, more than 229,000 individuals have participated in HUNT [16].

For HUNT-ADAPT, the principal clinical and biomarker framework is based on HUNT4 70+ and the subsequent 4-year AiT follow-up, while earlier HUNT waves are used to characterize cumulative and trajectory-based exposures across adulthood and later life. The overall study design and analytic framework are shown in Figure 1.

**Figure 1.**
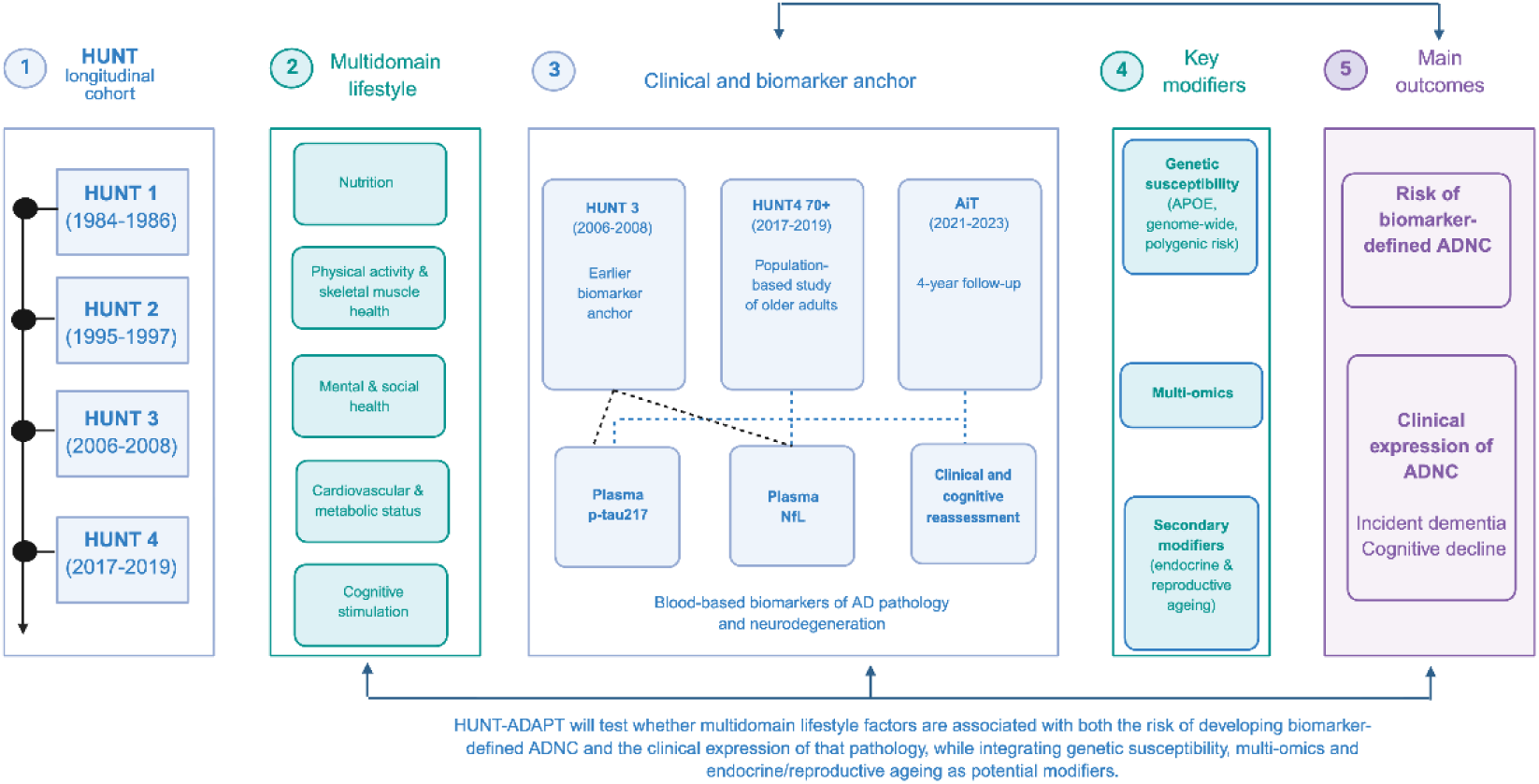
Conceptual framework and study design of the HUNT-ADAPT study. Created with BioRender. The HUNT-ADAPT study integrates longitudinal multidomain lifestyle and health-related data collected across the HUNT waves (HUNT1–HUNT4) to examine their association with biomarker-defined Alzheimer disease neuropathologic change (ADNC) and its clinical expression. Five multidomain lifestyle domains are evaluated: nutrition, physical activity and skeletal muscle health, mental and social health, cardiovascular and metabolic status, and cognitive stimulation. Biomarker anchoring is based on plasma p-tau217 and NfL measurements obtained in HUNT3 and HUNT4 70+, with approximately 4-year follow-up and clinical/cognitive reassessment in the AiT study. Genetic susceptibility, multi-omics data, and endocrine/reproductive ageing are incorporated as potential modifiers of pathology risk and clinical vulnerability. The primary outcomes are biomarker-defined ADNC and the clinical expression of ADNC, including incident dementia and cognitive decline. **Abbreviations:** ADNC, Alzheimer disease neuropathologic change; AiT, Ageing in Trøndelag; HUNT, Trøndelag Health Study; p-tau217, phosphorylated tau 217; NfL, neurofilament light chain.

### 2.2. Source population and study waves

HUNT is an open, prospective cohort study in which all adult residents of Trøndelag County aged 20 years or older are invited to repeated health surveys. Four major waves have been completed: HUNT1 (1984–1986), HUNT2 (1995–1997), HUNT3 (2006–2008), and HUNT4 (2017–2019). Participation rates were 89.4% in HUNT1, 69.5% in HUNT2, 54.1% in HUNT3, and 54.0% in HUNT4 [17]. See table 1.

**Table 1.** Overview of HUNT waves and participant numbers.

| **HUNT wave / sub-study** | **Years** | **Participants (N)** |
| --- | --- | --- |
| HUNT1 | 1984–1986 | 77,212 |
| HUNT2 | 1995–1997 | 65,237 |
| HUNT3 | 2006–2008 | 50,807 |
| HUNT4 | 2017–2019 | 56,078 |
| HUNT4 70+ | 2017–2019 | 9,956 |
| AiT | 2021–2023 | 5,729 |

**Abbreviation:** AiT, Ageing in Trøndelag.

In the original HUNT catchment area, 19,403 residents aged 70 years or older were eligible and invited to the HUNT4 70+ sub-study; 9,956 participated (51.3%), and 8,949 provided blood samples. Standardized cognitive, physical, blood sampling, and clinical assessments were performed at field stations, private homes, or nursing homes to maximize participation among frail older adults. Participants were subsequently invited to the AiT follow-up (2021–2023), in which 5,729 underwent repeat questionnaires, clinical assessments, cognitive testing and blood sampling.

HUNT4 70+ and AiT provide the core biomarker and clinical framework, whereas earlier HUNT waves provide repeated exposure data across adulthood. Because the youngest HUNT4 70+ participants were approximately 40 years old at HUNT1, HUNT-ADAPT can capture up to four decades of adult-life exposure history before late-life biomarker and cognitive assessment.

Genome-wide genetic data are available in more than 99% of HUNT4 70+ participants. In addition, HUNT4 subsets include fecal metagenomic sequencing [18] data and large-scale proteomic data, including SomaScan 11K profiling in a substantial HUNT4 70+ subset. These resources support analyses of genetic susceptibility, host–microbiome interactions, and broader multi-omics signatures relevant to ADNC risk and clinical expression.

National registry linkage will complement HUNT data through the Norwegian personal identification number. For the initial HUNT-ADAPT analyses, linkage will be limited to the Norwegian Prescribed Drug Registry and Norwegian Cause of Death Registry to ascertain medication use, mortality status and date of death. Linkage to additional national healthcare registries may be considered in subsequent analyses where directly required by specific research questions and subject to separate approvals.

### 2.4. Temporal framework and analytic samples

HUNT-ADAPT is conceived as a long-term course research program integrating repeated exposure assessments across the HUNT study period, from HUNT1 to HUNT4/AiT. The temporal framework will vary according to the research question and the availability of comparable measures across waves. For analyses of late-life biomarker status and subsequent cognitive or clinical outcomes, HUNT4 70+ will serve as the main analytic reference point, with follow-up based on the 4-year AiT reassessment. Earlier HUNT waves will be used where relevant to characterize cumulative or trajectory-based exposures across adulthood and later life, particularly for physical activity, cardiometabolic health, mental and social health, cognitive stimulation, and other repeated lifestyle-related measures. For variables available only in late life, analyses will rely primarily on HUNT4 70+ and AiT. The temporal structure of the HUNT-ADAPT data sources, including blood-based biomarker availability and cognitive/clinical assessments, is summarized in Figure 2.

**Figure 2.**
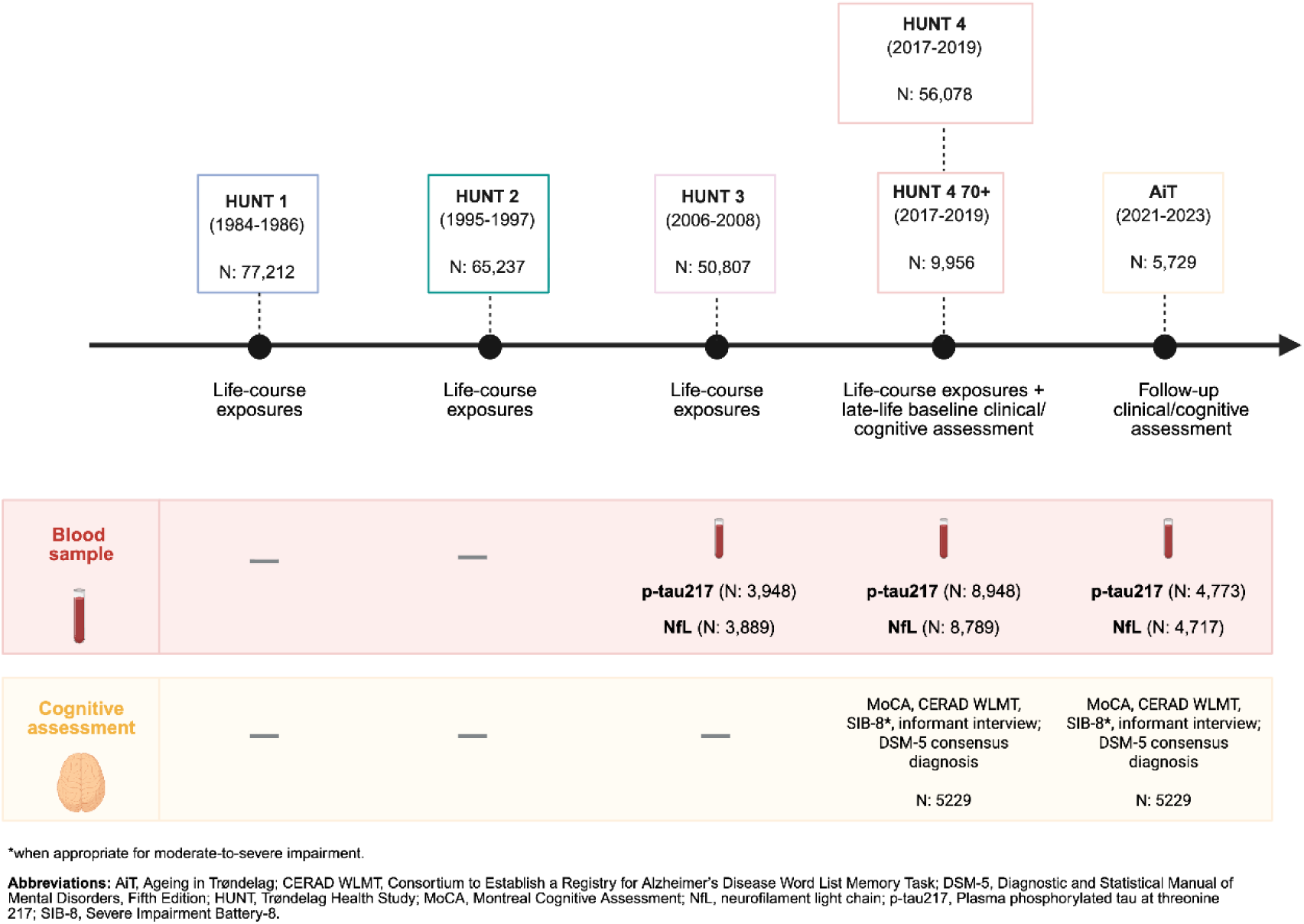
Timeline of HUNT-ADAPT data sources, blood-based biomarkers, and cognitive assessments. Created with BioRender. The figure illustrates the longitudinal structure of the HUNT and the nested HUNT4 70+ and AiT cohorts used in HUNT-ADAPT. The timeline includes HUNT1 (1984–1986), HUNT2 (1995–1997), HUNT3 (2006–2008), HUNT4 (2017–2019), HUNT4 70+, and the follow-up AiT assessment (2021–2023). Participant numbers for each wave are shown. Blood-based biomarkers of Alzheimer disease pathology and neurodegeneration, including p-tau217 and NfL, were available in subsets of participants from HUNT3, HUNT4 70+, and AiT. Cognitive and clinical assessments performed in HUNT4 70+ and AiT included the MoCA, CERAD WLMT, SIB-8, informant interviews, and DSM-5 consensus-based diagnostic evaluations.

The study population includes the full HUNT4 70+ cohort (n = 9,956), of whom 5,729 also participated in AiT. AD biomarkers have been analyzed, including p-tau217 (n = 3,948) and NfL (n = 3,889) in HUNT3; p-tau217 (n = 8,948) and NfL (n = 8,789) in HUNT4 70+; and p-tau217 (n = 4,773) and NfL (n = 4,717) in AiT. Final analytic sample sizes will vary according to the overlap and availability of exposures, biomarkers, covariates, modifiers, mediators, and outcomes across analyses.

The current protocol is centered on analyses based on HUNT1–HUNT4, HUNT4 70+, and AiT, while allowing future extensions through HUNT5, expected in 2028–2030, and continued registry-based follow-up, subject to data availability and regulatory approvals.

## 3. Multidomain lifestyle exposures

The main analysis will examine whether a multidomain lifestyle profile, operationalized through five FINGER-aligned domains (Table 2)—nutrition, physical activity and skeletal muscle health, mental and social health, cardiovascular/metabolic status, and cognitively stimulation activities—is associated with both the risk of biomarker-defined ADNC and the clinical expression of biomarker-defined pathology, including longitudinal test score change and incident dementia. Domain-specific scores will be constructed from harmonized, conceptually coherent HUNT measures summarized in Table 2 and fully detailed in Appendix Table A1. These domain-specific measures will enter the main analyses as a multidimensional lifestyle profile, with an overall composite lifestyle score considered in complementary or sensitivity analyses where conceptually and empirically justified [19].

**Table 2.** Summary of HUNT-ADAPT lifestyle domains and representative variables.

| **Domain** | **Representative variables** | **HUNT wave** |
| --- | --- | --- |
| **Nutrition** | Fruits/berries; vegetables; low-fat fish; high-fat fish; processed meat; sugar-sweetened drinks; juice; semi-whole-grain bread; whole-grain bread; extra-coarse whole meal bread | HUNT4 |
| **Physical Activity and Skeletal Muscle Health** | Exercise frequency; exercise intensity; exercise duration; composite physical activity volume; SPPB total score; gait speed; grip strength | HUNT1–HUNT4 |
| **Mental and Social Health** | HADS anxiety; HADS depression; HADS total/global distress; previous mental health problems; loneliness; neuropsychiatric symptoms | HUNT2–HUNT4 |
| **Cardiovascular and Metabolic Status** | Systolic blood pressure; diastolic blood pressure; body mass index; waist circumference; HbA1c; total cholesterol; triglycerides | HUNT1–HUNT4 |
| **Cognitive Stimulation** | Employment status; entertainment/leisure time; video games; social/friendship activities; knowledge-related activities; job-related activities; toileting; washing; bathing; dressing; eating | HUNT4 |

**Abbreviations:** HUNT, Trøndelag Health Study; HADS, Hospital Anxiety and Depression Scale; HbA1c, glycated hemoglobin; SPPB, Short Physical Performance Battery

### 3.1. Nutrition

Healthier dietary patterns, including Mediterranean and the Mediterranean-DASH Intervention for Neurodegenerative Delay (MIND) diet, have been associated with slower cognitive decline and lower dementia risk, whereas poorer-quality diets may contribute to adverse cardiometabolic and cognitive outcomes. Diet also shapes gut microbiome composition and function [20–22], with downstream effects on short-chain fatty acid production, vitamin metabolism, and inflammatory signaling, all of which may be relevant to neurodegeneration and resilience to pathology [23–26].

Within HUNT-ADAPT, the core dietary component will be derived from food frequency data, including measures of habitual intake of various food groups (e.g., fruits, vegetables, whole grains, fish/seafood, red and processed meats, dairy, dietary fats, candy and sugar-sweetened drinks). Dietary exposure will be operationalized as composite score based on HUNT questionnaire data on habitual food intake. Components will be scored according to direction of association with health, drawing on simplified diet scoring approaches previously applied in HUNT, such as LiFeCRC model by Brenne et al. (2024) [27], and summed to form a dietary domain within the multidomain lifestyle score.

### 3.2. Physical activity and skeletal muscle health

Physical activity and skeletal muscle health are closely linked to vascular health, metabolic regulation, inflammation, and functional reserve, and may therefore influence both the risk of biomarker-defined pathology and its clinical expression. The core physical activity component will be based on the combined physical activity index previously developed and applied in HUNT, which integrates self-reported frequency, intensity, and duration of leisure-time activity into a summary measure of weekly activity volume. Prior HUNT studies have used this index to characterize long-term physical activity patterns over time, supporting its use for cumulative and trajectory-based analyses across adulthood and later life [28–32].

Repeated physical activity measures across available HUNT waves will be used to derive the core long-term activity component, whereas late-life physical performance measures, including Short Physical Performance Battery (SPPB), gait speed, and grip strength, will be examined separately as markers of skeletal muscle health and functional reserve. These measures will not contribute directly to the core multidomain lifestyle score but will be examined in complementary domain-specific analyses.

### 3.3. Mental and social health

Depression, anxiety, loneliness and reduced social connectedness have been associated with poorer cognitive ageing and higher dementia risk [33, 34], potentially through behavioral, vascular, inflammatory, neuroendocrine, and psychosocial pathways [35–37]. HUNT data have also previously linked midlife mental distress to dementia risk over long-term follow-up [38]. However, late-life affective and social symptoms may also reflect prodromal disease, or a behavioral symptomatic profile, making temporality particularly important in this domain.

Depressive and anxiety symptoms will be assessed using the Hospital Anxiety and Depression Scale (HADS), including anxiety, depression, and total scores across HUNT waves. Additional indicators include self-reported mental health help-seeking and loneliness across HUNT waves. Self-reported symptoms will be complemented by informant-reported neuropsychiatric symptoms, including symptoms captured by the Neuropsychiatric Inventory (NPI) in AiT and, for a smaller subgroup mainly among participants with MCI, in HUNT4 70+, with a focus on indicators relevant to mild behavioral impairment [39]. These measures will be harmonized into mental and social health profiles and analyzed as midlife, late-life, cumulative, and trajectory-based exposures where possible, to address reverse causation and distinguish antecedent risk from potential prodromal symptoms. This domain will inform both the multidomain lifestyle score and domain-specific analyses.

### 3.4. Cardiovascular and metabolic status

Cardiovascular and metabolic status have been consistently linked to cognitive decline and dementia risk [40, 41], and HUNT data have previously supported life-course analyses of cardiometabolic and vascular risk trajectories [42]. This domain will include repeated measures of blood pressure, pulse, adiposity, including body mass index and waist–hip ratio, glycemic status, lipid profile, renal function, inflammatory markers, and self-reported cardiometabolic disease across HUNT waves. Blood pressure medication indicators will be used where appropriate to inform exposure classification or confounding control. Where repeated measures are available, analyses will distinguish midlife, late-life, cumulative, and trajectory-based profiles, recognizing that late-life cardiometabolic changes may reflect prodromal disease or competing health processes, as recently shown for adiposity in HUNT [43].

### 3.5. Cognitively stimulation activities

Cognitive stimulation is a potentially modifiable determinant of cognitive ageing and contributes to lower risk of cognitive impairment and dementia [12, 44]. Through mechanisms related to neuroplasticity, adaptive network reorganization, and cognitive reserve, cognitive stimulation may support the maintenance of cognitive function across ageing [45]. Education may provide an early foundation for cognitive reserve, occupational complexity may sustain cognitive engagement throughout adulthood, and leisure activities may contribute to continued cognitive enrichment in later life [8].

Within HUNT-ADAPT, this domain will be operationalized using available HUNT measures related to educational attainment, occupational complexity, and cognitively stimulating leisure activities. These measures will first be examined as separate domain-specific indicators reflecting early-life, midlife, and later-life sources of cognitive stimulation. They will subsequently be harmonized into a cognitive stimulation domain score, in which higher values indicate a more favorable cognitive stimulation profile across the life course.

## 4. Biomarkers and cognitive outcomes

### 4.1. Blood-based biomarkers

Plasma p-tau217 is the primary variable used to operationalize biomarker-defined ADNC. Plasma NfL will be examined as a complementary marker of neurodegeneration.

The plasma p-tau217 measurements used in HUNT-ADAPT are those generated for the HUNT4 70+ and HUNT3 samples in the source population [15]. Quantification was performed at the Clinical Neurochemistry Laboratory, University of Gothenburg, Sweden, on the Simoa HD-X platform (Quanterix) using the commercially available, previously validated ALZpath p-Tau 217 Advantage PLUS assay (Quanterix) for plasma p-tau217 and the Simoa NF-light assay (Quanterix) for plasma NfL, with samples analyzed between January and August 2024. Sample collection, processing, and storage followed standardized HUNT biobank procedures for EDTA plasma, with controlled pre-analytical handling, continuous −80 °C storage, and temperature-regulated dry-ice transport prior to analysis [14]. Analytical performance for plasma p-tau217 was monitored using calibrators run in duplicate, with outlier replicates excluded prior to calibration-curve fitting, and three quality-control levels of pooled human plasma run in duplicate at the start and end of each analytical run. Across the assayed range (mean concentrations of approximately 0.4, 1.5, and 1.9 pg/mL), repeatability (intra-assay %CV) was approximately 8.9–10.6% and intermediate precision (inter-assay %CV) approximately 10.3–15.9%.

Biomarker-defined ADNC is operationalized using a two-cutoff approach, consistent with recommendations from the Global CEO Initiative on Alzheimer’s Disease and with the externally derived and previously validated thresholds applied in the source cohort [4, 15, 46, 47]. A lower cutoff of 0.40 pg/mL (approximately 95% sensitivity) is used to rule out ADNC, and an upper cutoff of 0.63 pg/mL (approximately 95% specificity) is used to rule in ADNC; concentrations between these values define an intermediate zone [4]. The primary ADNC-positive classification in HUNT-ADAPT uses the 0.63 pg/mL threshold, consistent with the population p-tau217 positivity prevalence applied elsewhere in this protocol [15]. Plasma NfL will be classified using age-dependent cutoffs, with concentrations above 20 pg/mL at age 70 years and above 35 pg/mL at age ≥71 years indicating elevated neuroaxonal injury.

### 4.2. Cognitive outcomes

Clinical outcomes will include longitudinal cognitive test-score change and incident dementia. Cognitive change will be assessed using repeated cognitive measures from HUNT4 70+ and AiT, anchored in the Norwegian version of the Montreal Cognitive Assessment (MoCA). Additional cognitive characterization includes the Word List Memory Task (WLMT) from the Consortium to Establish a Registry for Alzheimer’s Disease (CERAD), administered to participants with MoCA scores ≥22 who recalled at least one word on the MoCA delayed-recall task, and the nine-item Meta-Memory Questionnaire (MMQ) for subjective memory performance. For nursing-home participants with suspected moderate-to-severe dementia, the Severe Impairment Battery-8 (SIB-8) was used instead of the MoCA. Global cognitive and functional impairment was assessed using the Clinical Dementia Rating (CDR) scale.

Incident dementia and MCI will be defined according to the established HUNT4 70+/AiT diagnostic framework described elsewhere [48].

## 5. Multi-omics

Longitudinal multi-omics resources within HUNT include genome-wide genotyping data from 88,517 individuals across HUNT2–4, nuclear magnetic resonance (NMR)-based blood metabolomics in large HUNT2/HUNT3 subsets, proteomic data in approximately 5,700 participants using SomaLogic and Olink platforms, and fecal metagenomic sequencing of the gut microbiome in approximately 13,268 HUNT4 participants [49]. The SomaLogic sample is largely selected from HUNT4 70+. Additional NMR-based metabolomics using the Nightingale Health platform and proximity extension assay (PEA)-based proteomics using Olink are available or will be generated in large HUNT plasma subsamples. For HUNT-ADAPT, the analytic sample size for each omics layer will depend on overlap with the HUNT4 70+ cohort, blood-based biomarker availability, AiT follow-up, and required covariates. These data will be used to identify biological signatures and pathways relevant to ADNC risk and pathology-related clinical outcomes.

### 5.1 Genetics

Genetic susceptibility will be examined using *APOE* ε2/ε3/ε4 genotype, with analyses considering *APOE* ε4 carrier status and, where sample size permits, ε4 allele dose. *APOE* ε4 is the strongest common genetic risk factor for late-onset AD [50]. This will be combined with genome-wide or polygenic risk measures to capture as much as possible of the genetic susceptibility across common and rare variants. Recent findings from largest GWAS to date shows that common variants explain over 15% of the variation, with polygenic risk score prediction performance of 17% in clinical samples [51]. This is biologically relevant because polygenic risk scores can capture additional genome-wide susceptibility beyond the *APOE* locus [52, 53] and AD polygenic risk has been associated with cognitive performance, dementia risk, and AD-related biomarker or neuropathological profiles [54, 55]. Genetic susceptibility will be incorporated as a covariate, stratification factor, and potential modifier in analyses of pathology risk and clinical expression.

### 5.2. Metabolomics

The Nightingale NMR metabolomics panel quantifies 249 metabolites, encompassing 228 lipids, lipoproteins or fatty acids, and 21 non-lipid traits, including amino acids, ketone bodies, fluid balance, glycolysis-related metabolites, and inflammation-related metabolites [56]. In recent work, 534 independent loci associated with metabolites were identified, explaining more than 70% of the genetic variance in fatty acid levels, greater than what is observed for most complex traits, in the largest metabolite quantitative trait loci study to date (n > 300,000) [57]. HUNT includes Nightingale NMR plasma metabolomics data from HUNT2 (n = 672) and HUNT3 (n = 19,921). Standard preprocessing pipelines, including the ukbnmr R package, will be used to remove sources of technical noise across cohorts [58].

### 5.3. Proteomics

The antibody-based Olink Explore 3072 PEA platform uses next-generation sequencing readout and quantifies up to 2,941 protein analytes. Unlike more traditional techniques, such as mass spectrometry-based proteomics, Olink’s PEA methodology allows quantification of less abundant proteins, including neurological, cardiometabolic, and inflammation-related proteins. HUNT includes plasma proteomics data using Olink Target 96 panels from HUNT2 and HUNT3 subsets, with proteins that directly overlap with the Olink Explore platform measured in UK Biobank [49]. In HUNT4 70+, SomaScan 11K proteomic profiling has also been performed in a large subset.

### 5.4. Microbiome

Stool microbiome data are available from the HUNT gut microbiome resource. In HUNT, 13,268 of 55,561 participants submitted stool samples. Stool was collected by participants at home using filter papers and sent to the HUNT Biobank, where samples were stored at-80°C. DNA was extracted from three 6-mm punched discs from each filter card after bead-beating, using the Microbiome MagMAX Ultra kit on the KingFisher Flex platform, and quantified using Quant-It dsDNA reagent. DNA fragmentation and library preparation were performed using the Celero EZ DNA-seq Core Module. Paired-end sequencing was performed on an Illumina NovaSeq 6000 platform at 2× 150 bp, with a mean sequencing depth of 22.9 million reads per samples [59, 60].

Metagenomic profiling was performed using the Clinical Microbiomics Human Microbiome Profiler (CHAMP) pipeline [61]. After host read removal, adapter and quality trimming, and post-sequencing quality control, 12,887 high quality HUNT samples were retained for microbiome profiling [62]. Microbiome profiling generated species-level microbial abundance estimated functional potential, viral abundance, and alpha and beta diversity measures. Current profiles were generated using the CHAMP pipeline and annotated according to Genome Taxonomy Database (GTDB) nomenclature.

For the present study, 2.933 participants have overlapping HUNT4 fecal microbiome and HUNT70+ data, defining the main microbiome analytic sample for analyses of AD-related pathology, neurodegeneration, and clinical vulnerability. Additional HUNT4-based overlap is available for 1,411 participants with fecal microbiome and HUNT 4 Diet data, and 1,252 participants with fecal microbiome and HUNT Dental Health data. Host–microbiome multi-omics data integration will be used where feasible to explore pathways linking multidomain lifestyle profiles to AD-related pathology, neurodegeneration, and clinical vulnerability [63, 64].

## 6. Reproductive, endocrine, and oral health ageing

Oral health will be examined as a complementary marker of ageing-related vulnerability, using available HUNT measures of dentition, chewing difficulties, dry mouth, and missing tooth surfaces. Emerging evidence suggests that oral disease and tooth loss may contribute to cognitive decline through systemic inflammation, nutritional impairment, and frailty-related pathways [65–67].

Reproductive and endocrine ageing indicators will be explored in parallel, including reproductive timing and history, menopausal hormone therapy, surgical reproductive history, and serum measures of sex and thyroid-related hormones. Additional hormonal markers may be incorporated if approved assays from stored biosamples become available. These variables will be analyzed as secondary ageing-related domains and will not contribute to the main multidomain lifestyle score [68, 69].

## 7. Covariates and potential confounders

All multivariable analyses will adjust for prespecified confounders selected a priori according to the specific research question, informed by directed acyclic graphs. Candidate covariates include age, sex, education, cognitive performance, *APOE* and broader genetic susceptibility, vascular and metabolic comorbidity, neuropsychiatric and behavioral factors, smoking, alcohol use, sensory impairment, traumatic brain injury, and other relevant health-related conditions, with final subsets defined separately for each research question. Because some lifestyle-related, psychosocial, and cardiometabolic variables may function either as components of domain-specific scores or as covariates depending on the analysis, their analytical role will be determined according to a prespecified conceptual framework. The study also explicitly acknowledges time-varying exposure and time-varying confounding across the life course.

## 8. Statistical analysis

The analytical strategy is structured around two related questions: whether multidomain lifestyle profiles are associated with the risk of biomarker-defined ADNC, and whether they modify the clinical expression of ADNC once pathology is present. The primary exposure framework will be the FINGER-aligned multidomain lifestyle profile, represented by harmonized domain-specific measures. Domain-specific measures will be modelled jointly where appropriate, with selected individual components and, where conceptually and empirically justified, an overall composite lifestyle score examined in complementary or sensitivity analyses. For the first question, biomarker-defined ADNC, operationalized primarily using plasma p-tau217, serves as the outcome. For the second question, biomarker-defined ADNC serves as the main pathological anchor, and the primary clinical outcomes will be longitudinal cognitive change from HUNT4 70+ to AiT and incident dementia at the AiT reassessment.

Analyses of biomarker-defined ADNC risk will be cross-sectional within HUNT4 70+. Plasma p-tau217 will be modeled both as a continuous outcome, using linear regression, and as a binary positivity classification at the previously validated 0.63 pg/mL cutoff, using logistic regression. These models will estimate whether the multidomain lifestyle profile, domain-specific exposures, and selected individual components are associated with the risk of biomarker-defined ADNC.

For analyses based on HUNT4 70+ and AiT, cognitive change will be modeled using HUNT4 70+ cognitive performance and follow-up interval as model terms. The HUNT4 70+ dementia assessment will first be used to define dementia status at late-life assessment. Among participants without dementia at HUNT4 70+, incident dementia will be analyzed as a 4-year cumulative clinical outcome at AiT, defined as transition from no dementia at HUNT4 70+ to dementia at follow-up according to the established consensus DSM-5 diagnostic framework [48]. Because the exact date of dementia onset is not observed, time-to-event analyses will be secondary or exploratory, with death handled as a competing event when relevant.

A central aim of HUNT-ADAPT is to assess whether multidomain lifestyle profiles modify the clinical expression of biomarker-defined ADNC. Interaction analyses will test whether the association between ADNC and clinical outcomes differs according to the multidomain lifestyle profile, with complementary analyses of selected domain-specific exposures and, where justified, an overall composite lifestyle score. For binary clinical outcomes, the primary scale of inference will be additive, with emphasis on absolute risk differences; multiplicative interaction will be reported as a complementary measure. For continuous cognitive outcomes, interaction effects will be interpreted as differences in predicted cognitive change across levels of ADNC and lifestyle exposure.

All multivariable analyses will adjust for prespecified confounders selected a priori according to the specific research question. Available HUNT medication indicators will be considered, where relevant, for exposure classification, confounding control, or sensitivity analyses. Missing data patterns will be examined, and multiple imputation will be used when justified by the extent and structure of missingness. Inverse probability weighting will be considered to address differential participation, biomarker availability, and follow-up attrition. Weighting models will build on published HUNT weighting strategies and incorporate variables associated with participation and sample availability, including age, sex, education, cognitive status, *APOE* ε4 status, and relevant health characteristics.

Sensitivity and complementary analyses will assess the robustness of findings to alternative p-tau217 cut-offs, continuous p-tau217 measures, complementary NfL analyses, alternative exposure specifications, baseline heterogeneity, regression to the mean, baseline-dependent change in repeated late-life outcome analyses, and alternative approaches to missing data and selection. These analyses may include stratification by baseline cognitive level or diagnostic status, restriction analyses excluding participants with advanced baseline impairment, comparison of alternative longitudinal model specifications, and analyses with and without inverse probability weights.

### 8.1. Power and sample size

Final analytic sample sizes will vary across analyses according to the overlap of exposures, biomarkers, covariates, modifiers, and outcomes. The planned analyses fall into three main analytic families with distinct sample size and precision structures: analyses of biomarker-defined ADNC, analyses of pathology-related cognitive change, and analyses of incident clinical dementia. The scenarios below are intended as illustrative detectable-effect estimates under conventional assumptions of 80% power and a two-sided 5% significance level.

#### Lifestyle and risk factors as determinants of biomarker-defined ADNC

The first family of analyses asks whether multidomain lifestyle and individual risk factors are associated with biomarker-defined ADNC at the time of HUNT4 70+. Plasma p-tau217 is analyzed both as a continuous outcome and as a binary positivity classification, using the previously validated 0.63 pg/mL cutoff with an expected positivity prevalence of approximately 33% [15]. The available sample is up to approximately 9,500 HUNT4 70+ participants, with n = 8,948 having plasma p-tau217 (mean 0.58 pg/mL, SD 0.48 pg/mL). As an illustrative scenario, comparing a risk-factor-exposed subgroup of approximately 14% of the sample (a prevalence comparable to that of diabetes in HUNT) with the remainder, the design provides approximately 80% power to detect a difference in mean p-tau217 of about 0.04 pg/mL and approximately 98% power to detect a difference of 0.06 pg/mL, the latter corresponding to roughly five years of biological ageing in HUNT. For continuous exposures such as physical activity, statistical power is generally greater than for the equivalent dichotomized comparison. For binary p-tau217 positivity, the design supports detection of odds ratios on the order of 1.07 per standard deviation of a multidomain lifestyle exposure measure at 80% power, with larger effects required for individual domain components and for stratified subgroup analyses.

#### Effect modification of biomarker-defined ADNC on longitudinal cognitive change

The second family of analyses asks whether multidomain lifestyle exposures modify the association between biomarker-defined ADNC and cognitive status at HUNT4 70+, as well as the association between baseline biomarker-defined ADNC and cognitive change at the AiT reassessment. The broad benchmark for the AiT-reassessed sample with valid follow-up is approximately n = 5,229, with the effective analytic sample further constrained by overlapping availability of baseline p-tau217 in HUNT4 70+, complete baseline and follow-up cognitive data, and required covariates. The estimand of interest is the interaction between baseline p-tau217 (continuous or dichotomized) and the multidomain lifestyle exposure in models of cognitive change conditional on baseline cognitive performance. Because interaction effects are estimated with lower precision than the corresponding main effects, this family represents the more constrained scenario for statistical power even with several thousand participants. Under conventional assumptions, the design is expected to support detection of interaction effects in the small-to-moderate range. Using Cohen’s conventions for the incremental variance explained by the interaction term, this corresponds approximately to f² of 0.02 (small) and 0.15 (moderate).

### 8.2. Estimating causal effects of lifestyle on ADNC - a Target Trial Emulation (TTE)

As a secondary analysis, HUNT-ADAPT will estimate the causal effect of prespecified changes in modifiable lifestyle components on ADNC detected at follow-up. In line with the multidimensional lifestyle framework, the TTE will define interventions on selected harmonizable lifestyle components, considered separately and jointly. For that purpose, we will emulate a hypothetical target trial nested within HUNT4 70+ among participants free of clinical dementia and without ADNC at baseline. AiT will serve as the fixed follow-up assessment approximately four years later.

The primary endpoint will be ADNC detected at AiT (yes/no). Because death before AiT precludes observation of ADNC at follow-up, the primary estimand will be defined as the counterfactual absolute difference in the four-year risk of ADNC detection before death under a prespecified lifestyle-improvement policy versus the observed lifestyle. That means, at baseline participants with current smoking are counterfactually shifted to non-current smoking; participants below the physical activity threshold are shifted to the minimum activity category; participants with high-risk alcohol consumption are shifted to low-risk/moderate consumption. Note that such counterfactual shifts can be performed separately for each lifestyle component and also jointly, hence mimicking a multicomponent lifestyle intervention. Participants already meeting a favorable criterion remain unchanged.

In sensitivity analyses, we will consider alternative endpoint definitions, including a composite endpoint of ADNC or death before AiT and analyses using inverse-probability-of-censoring adjustment for non-participation or missing AiT assessment.

The primary analysis will use targeted maximum likelihood estimation for a fixed-time endpoint, incorporating flexible machine-learning models for the outcome, exposure, and, where applicable, censoring or assessment mechanisms. Inverse probability of treatment weighting (IPTW) and outcome-regression/g-computation analyses will be used as comparative estimators. We will assess the plausibility of causal identification assumptions, including exchangeability, consistency, positivity, and independent censoring/assessment conditional on measured baseline covariates. Because only one major follow-up assessment is currently available after HUNT4 70+, the target trial will be formulated as a temporally constant baseline intervention on lifestyle profile rather than as a time-varying intervention. Further methodological details will be provided in the Appendix and in dedicated analysis protocols.

As a complementary life-course analysis, we will assess whether a restricted set of lifestyles and cardiometabolic factors (e.g., smoking, physical activity, BMI/obesity, hypertension, diabetes) measured repeatedly across HUNT waves can support a longitudinal target-trial emulation of dementia status at HUNT4 70+. Participants entering HUNT1 at age 40 years or older will be assumed to be free of dementia at baseline. Because not all lifestyle domains are assessed consistently from HUNT1 to HUNT4, the TMLE analysis will be limited to harmonizable domains such as smoking, physical activity, BMI, blood pressure/hypertension, diabetes, and possibly alcohol. Further details on the TTE framework are provided in the Appendix, with the conceptual causal structure illustrated in Figure A1.

## 9. Ethics, data access, and governance

This project will be based on existing HUNT data and approved linkage to Norwegian registries within the established HUNT consent and governance framework. Participation in HUNT is based on written informed consent for research use, with proxy consent used where relevant according to HUNT procedures and applicable regulations. The study will be conducted following approval by the Regional Committee for Medical and Health Research Ethics in Norway (REK). Analyses will mainly be limited to already available questionnaires, clinical, biomarker, genetic, microbiome, and approved registry-linked data within the scope of participant consent. Data will be handled within secure HUNT-approved infrastructures, with access restricted to approved researchers and in accordance with applicable data protection regulations.

Applications for access to HUNT Databank data will be submitted in accordance with procedures established by the HUNT Research Centre and the Norwegian University of Science and Technology (NTNU). The application will include a detailed project description, study aims, requested variables, data management plan, and relevant ethical approvals. Data access will be contingent on approval from the relevant ethical and data protection authorities and the HUNT Data Access Committee. All analyses will comply with applicable regulations for privacy, data security, and responsible handling of sensitive health information.

## 10. User Involvement

User involvement is a cornerstone of health research. It enhances the quality and relevance of research and expedites the implementation of findings. This project is guided by user involvement from WiseAge, a platform established at the Centre for Age-Related Medicine (SESAM), Stavanger University Hospital, a decade ago. The WiseAge platform harnesses the invaluable insights of the older generation, recognizing their unique perspective in a world marked by rapid demographic and technological shifts. With a diverse membership from the 60+ age group, WiseAge ensures a broad representation. The user panel and advisory board of WiseAge rigorously evaluated the study, and questioned the research staff about background, rationale and methods, offering critical insights. Dedicated user representatives have been allocated to the project and collaborate closely with researchers throughout the project, including the dissemination.

## 11. Discussion

HUNT-ADAPT is designed to test whether specific multidomain lifestyle profiles are associated with both the risk of biomarker-defined ADNC and the clinical expression of that pathology. The central hypothesis is that favorable multidomain profiles may be associated with lower probability of biomarker-defined pathology and, once pathology is present, attenuated clinical expression, including better cognitive outcomes, lower incident dementia risk, and greater clinical resilience. By integrating repeated life-course exposure data, blood-based biomarkers, genetic susceptibility, multi-omics resources, and late-life clinical phenotyping, HUNT-ADAPT aims to provide a more precise framework for understanding heterogeneity in pathology-related clinical outcomes.

Recent evidence supports a multidomain rather than single-factor approach to ADNC risk and age-related cognitive decline. A large multinational study across 34 countries showed that aggregated physical and social exposome models explained substantially more variance in brain ageing than individual exposures alone, supporting the view that brain health is shaped by interacting exposures rather than isolated risk factors [70]. HUNT-ADAPT extends this logic by examining multidomain lifestyle as a FINGER-aligned profile and through individual domains, while distinguishing associations with biomarker-defined pathology from associations with clinical outcomes.

The study is also well aligned with recent work on cognitive resilience, which has emphasized that preserved cognition in later life is unlikely to reflect a single protective factor and instead emerges from interacting biological, clinical, and environmental determinants [71]. By incorporating multi-omics, APOE genotype, broader genetic susceptibility, and endocrine/reproductive ageing, HUNT-ADAPT is positioned to examine why individuals with apparently similar pathological burden may follow different cognitive and clinical trajectories, including sex-specific patterns.

### 11.1. Strengths

The major strength of the project is the structure of HUNT. Few population-based cohorts combine this sample size, long follow-up, repeated life-course measures, standardized assessments, late-life blood-based biomarkers, extensive genetic and microbiome data, registry linkage, and longitudinal clinical phenotyping within a single framework. HUNT-ADAPT can capture up to four decades of adult-life exposure history before late-life biomarker and cognitive assessment, enabling multidomain lifestyle to be examined not only as a late-life exposure, but also as a cumulative and potentially trajectory-based profile across the life course. This design allows the study to address both biomarker-defined pathology risk and heterogeneity in its clinical expression within the same population-based framework.

Another major strength is its domain-specific multidomain structure, which moves beyond a generic lifestyle framework to focus on FINGER-aligned domains: nutrition, physical activity, mental and social wellbeing, cardiovascular/metabolic health, and cognitive stimulation. These exposures are unlikely to operate through identical mechanisms, may contribute differently to pathology risk and clinical expression, and allow comparison with previous multidomain prevention trials. Although pathways overlap, cardiovascular/metabolic status captures vascular and metabolic mechanisms; physical activity, functional and systemic reserve; nutrition, inflammatory and gut-related pathways; mental and social health, psychosocial, behavioral, and neuroendocrine mechanisms; and cognitive stimulation, reserve-related pathways. This domain-specific strategy is more informative than treating lifestyle as a single undifferentiated exposure.

### 11.2. Limitations

Several limitations should be acknowledged. First, despite the target trial emulation component, HUNT-ADAPT remains observational, and residual confounding, selection bias, reverse causation, and immortal time-related biases cannot be excluded. Second, plasma p-tau217 is an indirect blood-based proxy for AD pathology and does not replace amyloid/tau PET, cerebrospinal fluid biomarkers, or neuropathological confirmation, although it has shown strong diagnostic performance against established reference standards. Third, temporal coverage and harmonization will differ across domains, potentially limiting comparability of domain scores. Fourth, several components rely on self-reported information and may be vulnerable to misclassification; medication use is also incompletely captured in HUNT and will require cautious interpretation until prescription-registry data are available. Fifth, attrition, dropout, long intervals between cognitive assessments, death as a competing risk, and limited cognitive assessment before age 70 may influence longitudinal analyses. Sixth, effective sample size will vary according to overlap in exposures, biomarkers, modifiers, and outcomes. Finally, even within a life-course framework, it may remain difficult to disentangle whether multidomain lifestyle profiles are associated primarily with lower pathology risk, weaker clinical expression once pathology is present, or both.

## Conclusions

Overall, HUNT-ADAPT is intended to support a more precise and biologically grounded prevention framework in which specific multidomain lifestyle profiles are examined in relation to both biomarker-defined AD pathology and clinical heterogeneity. The study may help clarify whether specific multidomain lifestyle profiles are associated with lower probability of biomarker-defined pathology, attenuated clinical expression among individuals with pathology, or both, and thereby strengthen the conceptual basis for prevention and resilience research in AD.

## CRediT authorship contribution statement

Kevin O’Hara-Veintimilla: Conceptualization, Methodology, Project administration, Writing – original draft, Writing – review & editing, Visualization. Elisabeth Lind Melbye: Conceptualization, Methodology, Project administration, Writing – original draft, Writing – review & editing. Miguel German Borda: Conceptualization, Methodology, Writing – review & editing. Poppy AC Mallinson: Methodology, Writing – review & editing. Anita Lenora Sunde: Resources, Methodology, Writing – review & editing. Antoine Leuzy: Methodology, Writing – review & editing. Mark van der Giezen: Methodology, Writing – review & editing. Pier-Giorgio Masci: Methodology, Writing – review & editing. Yohannes Seyoum: Methodology, Writing – review & editing. Beatriz Pozuelo Moyano: Methodology, Writing – review & editing. Felipe Botero-Rodriguez: Methodology, Data curation, Writing – review & editing. Michael C. Craig: Methodology, Writing – review & editing. Lizhi Guo: Methodology, Writing – review & editing. Lingfeng Xue: Methodology, Writing – review & editing. Håvard K. Skjellegrind: Methodology, Writing – review & editing. Ragnhild Oesterhus: Resources, Methodology, Writing – review & editing. Audun Osland Vik-Mo: Conceptualization, Methodology, Supervision, Writing – review & editing. Diego A. Tovar-Rios: Methodology, Writing – review & editing. Mira G.P. Zuidgeest: Methodology, Writing – review & editing. Holger Fröhlich: Methodology, Supervision, Writing – review & editing. Chiara de Lucia: Methodology, Writing – review & editing. Richard Siow: Methodology, Writing – review & editing. Miia Kivipelto: Conceptualization, Methodology, Supervision, Writing – review & editing. Ole A. Andreassen: Conceptualization, Methodology, Resources, Supervision, Writing – review & editing. Geir Selbæk: Conceptualization, Methodology, Resources, Supervision, Writing – review & editing. Dag Aarsland: Conceptualization, Methodology, Project administration, Supervision, Writing – original draft, Writing – review & editing.

## Acknowledgment

The Trøndelag Health Study (HUNT) is a collaboration between HUNT Research Centre, Faculty of Medicine and Health Sciences, Norwegian University of Science and Technology (NTNU), Trøndelag County Council, Central Norway Regional Health Authority, and the Norwegian Institute of Public Health. We thank the population of Trøndelag for their willingness to contribute important data and biological material, and the staff at HUNT Research Centre and collaborating institutions for their contributions to the planning, collection, storage, and management of HUNT data and biological material. The Norwegian National Centre for Ageing and Health contributed to the funding of the HUNT4 70+ survey and funded the Ageing in Trøndelag (AiT) study. The HUNT4 70+ survey received additional funding from the Center for Oral Health Services and Research, Trondheim. The genetic investigations of the HUNT Study are a collaboration between researchers from the K.G. Jebsen Center for Genetic Epidemiology, NTNU, and the University of Michigan, with SNP genotyping performed by the Genomics Core Facility at NTNU.

