## Supplemental material for "Multidomain Lifestyle Profiles, Biomarker-Defined Alzheimer’s Pathology, and Clinical Expression: Design of the HUNT-ADAPT Study"

### Appendix

#### Target trial emulation (TTE)

TTE will be conducted in HUNT4 70+ among participants free of clinical dementia and without ADNC at baseline. AiT will serve as the fixed follow-up assessment approximately four years later. Because ADNC is assessed at discrete visits rather than continuously over time, the primary analysis will be formulated as a fixed-horizon TTE. The primary endpoint will be ADNC detected at AiT, with death before AiT handled explicitly because it precludes observation of ADNC at follow-up.

The primary intervention will be defined as a baseline modified treatment policy operating on modifiable HUNT4 lifestyle components. Unfavorable levels of selected components, such as physical activity, smoking, alcohol consumption, and sleep, will be counterfactually shifted to the nearest predefined favorable category, while favorable components will remain unchanged. This counterfactual intervention will be performed per individual lifestyle component and also jointly. Diet will be included if a robust HUNT4 diet-quality proxy can be constructed; cognitive and social engagement variables will be evaluated in sensitivity analyses. The primary estimand will be the counterfactual absolute difference in four-year ADNC detection risk before death under this (multicomponent) lifestyle-improvement policy compared with the observed lifestyle distribution.

The primary effect will be estimated using TMLE for a fixed-time endpoint, with flexible machine-learning models for the outcome regression and intervention/censoring mechanisms. Baseline demographic, clinical, genetic, cognitive, historical, and health-status covariates, including information from prior HUNT waves, will be used for adjustment according to the assumed causal structure. IPTW and g-computation analyses will be performed as comparative estimators. Cross-fitting will be used for robust nuisance-parameter estimation.

Because the primary endpoint is ADNC status at AiT, survival models for continuously observed ADNC onset will not be used as the primary modeling strategy. Sensitivity analyses will consider alternative endpoint definitions, including a composite endpoint of ADNC or death, inverse-probability-of-censoring adjustment for missing AiT assessment, and competing-risk-aware formulations of ADNC detection before death. Positivity, consistency, conditional exchangeability, and independent censoring/assessment assumptions will be evaluated using overlap diagnostics, weight distributions, covariate balance, and sensitivity analyses for unmeasured confounding.

As a complementary life-course analysis, we will evaluate whether a longitudinal target trial can be emulated among participants who were approximately 40-50 years old at HUNT1 and followed through subsequent HUNT waves. If lifestyle and time-varying confounder measurements can be harmonized sufficiently, we will use Longitudinal TMLE to estimate the effect of a repeated modified treatment policy on the cumulative risk of clinically detected dementia by late-life follow-up. Because dementia onset is not continuously observed, the endpoint will be treated as interval-censored or discretely observed at available assessment windows, rather than as exact time to onset. Death before dementia detection will be handled explicitly as a competing event or in sensitivity analyses using composite endpoints. ADNC detected in late-life assessed participants will be evaluated as a key secondary biomarker endpoint, with sensitivity analyses for survival and participation selection.

**Fig. A1.** Conceptual causal framework for time-varying exposure, confounding, censoring, and outcomes in the HUNT-MAP target trial emulation.

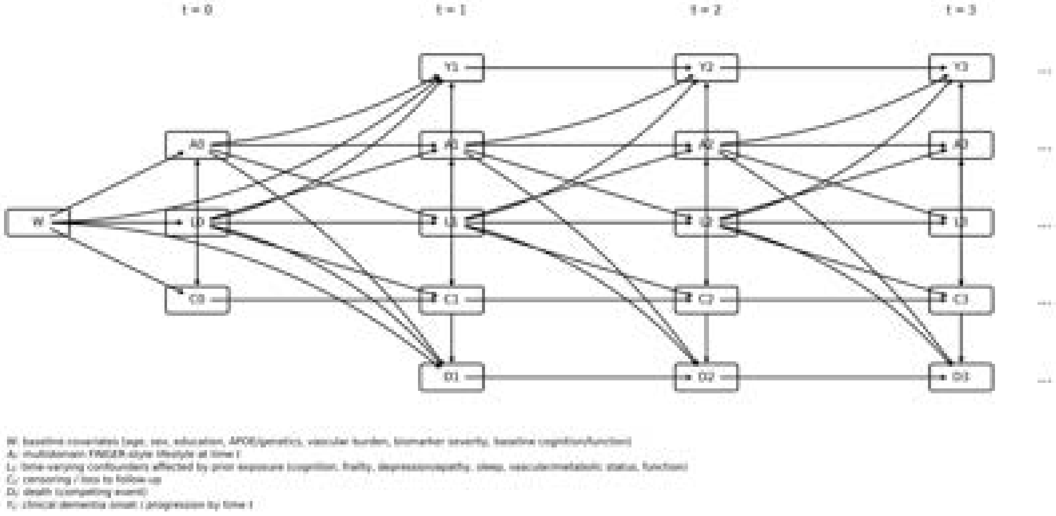

**Table A1**: Overview of multidomain variables included in the HUNT-MAP study

| Domain | Subdomain | Representative variables | HUNT wave |
|---|---|---|---|
| **Nutrition** | **Dietary habits and food intake** | Fruits/berries; vegetables; low-fat and high-fat fish; red/white meat; processed meat; sugary drinks; juice/smoothies; milk/yoghurt/dairy intake; whole-grain bread; meal frequency; alcohol intake; dietary supplements. | HUNT2–HUNT4 |
| | **Fatty acid biomarkers** | Palmitic acid; oleic acid; linoleic acid; alpha-linolenic acid; EPA; DHA; total omega-3 and omega-6 fatty acids | HUNT1 |
| | **Nutritional biomarkers** | Folate; albumin; fasting glucose; cholesterol; HDL-cholesterol | HUNT1–HUNT4 |
| | **Body composition and nutritional status** | Body fat mass; skeletal muscle mass; visceral fat; food intake; poor appetite | HUNT4 |
| **Physical Activity and Skeletal Muscle Health** | **Physical activity** | Exercise frequency; intensity; duration; physical activity index | HUNT1–HUNT4 |
| | **Physical performance** | SPPB; gait speed; grip strength | HUNT2–HUNT4 |
| | **Bone health and anthropometry** | Bone mineral density measures; T-scores/Z-scores; hip axis length; weight; upper arm circumference | HUNT4 |
| **Mental and Social Health** | **Affective symptoms** | HADS anxiety; HADS depression; global affective distress | HUNT2–HUNT4 |

| Domain | Subdomain | Representative variables | HUNT wave |
|---|---|---|---|
| | **Mental health history and social well-being** | Previous mental health problems; loneliness | HUNT2–HUNT4 |
| | **Neuropsychiatric symptoms** | Delusions; hallucinations; agitation; depression; anxiety; apathy; behavioral severity measures; empathy reduction; disinhibition; stereotypical behavior; eating behavior changes | HUNT4 |
| **Cardiovascular and Metabolic Status** | **Blood pressure and adiposity** | Systolic blood pressure; diastolic blood pressure; body mass index; waist circumference | HUNT1–HUNT4 |
| | **Glycaemic and lipid metabolism** | HbA1c; fasting/non-fasting glucose; diabetes history | HUNT1–HUNT4 |
| | **Lipid metabolism** | Total cholesterol; HDL-cholesterol; LDL-cholesterol; triglycerides; apolipoproteins; cholesterol ratios | HUNT2–HUNT4 |
| | **Renal, inflammatory, and cardiac biomarkers** | Creatinine; estimated glomerular filtration rate; micro C-reactive protein; interleukins; TNF-alpha; high-sensitivity troponin I | HUNT2–HUNT4 |
| | **Cardiovascular disease history and treatment** | Myocardial infarction; angina; stroke/brain hemorrhage; antihypertensive medication | HUNT1–HUNT4 |

| Domain | Subdomain | Representative variables | HUNT wave |
| --- | --- | --- | --- |
| | **Vascular lifestyle risk factors** | Smoking status; alcohol consumption | HUNT1–HUNT4 |
| **Cognitive Stimulation** | **Employment and leisure activities** | Employment; entertainment activities; friendship/social activities; knowledge-related activities; job-related activities; video games | HUNT4 |
| | **Functional independence and mobility** | Basic and instrumental activities of daily living; indoor walking; walking aids; driving license/current driving; cooking; housework; shopping; paying bills; medication management; going out; public transport use | HUNT4 |
| **Others** | **Oral health** | Dentition; chewing difficulties; dry mouth; missing tooth surfaces | HUNT4 |
| | **Reproductive and endocrine ageing** | Menopausal age; contraceptive and hormone therapy use; estrogen exposure; hysterectomy/oophorectomy; pregnancy history; thyroid hormones; sex hormone-binding globulin; testosterone | HUNT2–HUNT4 |

**Abbreviations:** HUNT, Trøndelag Health Study; DHA, docosahexaenoic acid; EPA, eicosapentaenoic acid; HADS, Hospital Anxiety and Depression Scale; HbA1c, glycated hemoglobin; IL, interleukin; HDL, high-density lipoprotein; LDL, low-density lipoprotein; SPPB, Short Physical Performance Battery; TNF, tumor necrosis factor.
